# CLINICAL VALIDATION OF SWAASA ARTIFICIAL INTELLIGENCE PLATFORM USING COUGH SOUNDS FOR SCREENING AND DIAGNOSIS OF RESPIRATORY DISEASES

**DOI:** 10.1101/2025.01.27.25321216

**Authors:** Balamugesh Thangakunam, Sujith Thomas Chandy, Gowrisree Rudraraju, Narayana Rao Sripada, Jayanthy Govindaraj, Charishma Gottipulla, Baswaraj Mamidgi, Shubha Deepti Palreddy, Nikhil kumar Reddy Bhoge, Harsha Vardhan Reddy Narreddy, P Prasanna Samuel, Devasahayam Jesudas Christopher, Manmohan Jain, Venkat Yechuri

**Affiliations:** Christian Medical College, Vellore,Tamil Nadu, 632004, India; Salcit Technologies, Kothaguda, Hyderabad, 500084, India

**Keywords:** Machine learning, Respiratory diseases, Cough sound characteristics, Airflow parameters, Respiratory conditions

## Abstract

**Introduction:** Analysis of cough sounds have the potential to give a clue regarding the underlying respiratory disease. The Swaasa AI platform using artificial intelligence technology can analyze the cough sounds and provides an output using cough as a marker. The Swaasa AI platform comes under the category of Software As a Medical Device (SaMD), the device can detect underlying respiratory condition as normal vs abnormal, the pattern of disease condition (obstructive, restrictive or normal), the severity of pattern (mild, moderate, severe) and presence of disease conditions for Asthma, COPD, ILD and Bronchiectasis.

**Methods:** Patients (age >= 18 years) presenting to our departments with respiratory symptoms were prospectively recruited into the study. The patient’s cough sound was recorded by a mobile device in which the Swaasa AI platform was installed. Spirometry was done and patients’ clinical diagnosis was taken as reference standard for comparison. Polychotomous variables were analyzed in binary form as normal and abnormal. Multi-class outcomes were defined as obstructive, restrictive, or normal pattern, mild-moderate-severe as pattern severity or disease conditions like Asthma, COPD, ILD and Bronchiectasis.

**Results:** We recruited 2179 patients with respiratory diseases and 827 normal individuals. Sixty-two percent of them were males. The Swaasa AI platform has 90% accuracy in distinguishing normal versus abnormal (having respiratory condition) as well as distinguishing normal from abnormal spirometry. The performance was consistent across demographic parameters such as age, gender, and BMI. The accuracy for multi-class outcomes were between 79%-84% for spirometry pattern and 76%-84.5% for pattern severity and 83%-93% for disease type classification.

**Conclusion:** Swaasa commends an elevated level of precision in classifying normal and abnormal conditions across demographic subgroups indicating its promise as a screening tool. While the accuracy measures emphasize further refinement to ensure consistent high performance across disease scenarios and severity levels, the study shows Swaasa’s reasonable potential in identification of specific respiratory diseases and their severity.

## INTRODUCTION

Respiratory sounds, produced during coughing and breathing, offer valuable insights for diagnosing respiratory diseases. Advances in technology now allow the acoustic characteristics of these sounds to be easily and non-invasively recorded, enhancing their effectiveness as diagnostic tools (1).

Recent advances in digital devices have led to the development of sophisticated technologies for recording and analyzing cough sounds. These technologies have been evaluated for screening, triaging, and evaluating treatment responses in patients with airway disorders like asthma and Chronic Obstructive Pulmonary Disease (COPD), as well as in those who had Covid-19 infection (2–6). Cough sound devices can indicate and analyze cough intensity, frequency, and decode cough signature for different respiratory conditions (23, 26–27).

Respiratory diseases encompass any issues in the respiratory system that may hinder normal lung function. In clinical practice, pulmonary function testing (PFT) provides crucial feedback on a patient’s lung status based on their pulmonary function parameters, aiding doctors in diagnosing respiratory diseases (7, 8). Spirometry is one of the most readily available and useful tests for pulmonary function.

Spirometry measures the volume of air exhaled at specific time points during complete exhalation by force, which is preceded by a maximal inhalation. The test requires patients to place their mouth on the spirometer, inhale deeply, and exhale forcefully to expel all air quickly. This process must be completed with maximum effort and repeated until three consistent measurements are obtained (9). Patients are instructed and guided through the test procedure by trained Respiratory therapists. Reference values for 1-second forced expiratory volume (FEV1), forced vital capacity (FVC), and the FEV1/FVC ratio are estimated based on the patient’s age, gender, and height. The ratios of the measured values to the reference values, expressed as FEV1% and FVC%, indicate the severity of respiratory disease (10). FEV1/FVC and FVC% help distinguish between obstructive, restrictive, and normal respiratory patterns, while FEV1% determines the severity of obstructive diseases (11).

In recent years, the use of home-based spirometry to monitor lung function in patients with Asthma, Chronic Obstructive Pulmonary Disease (COPD), interstitial lung disease (ILD) has gained attention in clinical practice and research (12–16). Home-based spirometry has the potential to increase convenience and accessibility for patients with Asthma, COPD, and ILD to improve the frequency of data collection, and make it easier for patients to receive regular assessments of their lung function. In addition, the integration of smartphone applications has facilitated communication and collaboration between patients and healthcare providers. With advances in machine learning (ML) and an increasing amount of health data available for analysis, it is becoming more feasible to use ML algorithms to improve both the quality and the interpretation of pulmonary function testing (18–19). Despite the potential benefits of using ML in home-based spirometry, most research has focused on automating current human tasks (e.g., diagnosis).

Traditional diagnostic tools for respiratory diseases such as spirometry; swab tests; bronchoscopies and radiological investigations like chest scans; X-rays are effective but often lack accessibility and affordability. This gap underscores the need for alternative solutions, where artificial intelligence (AI), particularly machine learning, emerges as a promising avenue, offering more accessible and cost-efficient diagnostic methods using devices as common as smartphones.

Our research investigates cough sound signal analysis, aiming to map these signals to their corresponding respiratory diseases and explore coughing as a potential indicator. We focus on identifying specific features within cough sounds for respiratory disease diagnosis, extending our study to differentiate cough signals in different respiratory diseases. On the other hand, recording cough sounds requires minimal training unlike what is needed for spirometry, no physical contact, and is effective for disease assessment (21, 12–16, 18–19). Clinical research shows that cough severity indicates respiratory disease progression (22).

Signal processing can extract diagnostic features from cough sounds, and studies have shown the importance of classifying these sounds to detect various respiratory illnesses (22). In this context, our previous studies have already proven strong correlation of cough sound characteristics with airflow characteristics including FEV1, FVC and their ratios, which are important in identifying the type of lung diseases (23). Following this, in the current study, we attempted to establish the high-level precision of the Swaasa AI platform (https://swaasa.ai/) in classifying normal and abnormal conditions across demographic subgroups, indicating it as a promise screening tool.

## METHODS AND MATERIALS

### Data collection

All patients included in the study were checked by a respiratory physician, followed by a spirometry and cough sound assessment using mobile phones on the same day, data collection process is illustrated in figure 1. Informed consent was obtained from all participants and/or their legal guardians. This was done to ensure that there was no delay between the index and reference tests. To record cough sounds, an in-built mobile microphone that is a part of the Swaasa AI platform was used. Five voluntary cough recordings were done per participant. Voluntary cough recording captures lung pathology by reflecting physiological changes in cough sound. Regardless of how cough is elicited (spontaneous or voluntary), the sensory receptors get triggered in case of a pathological condition. Each condition of the lung tends to produce a distinct sound profile. Since there were noise corrupted coughs, invalid coughs and void recordings, an average of 3 recorded coughs from each participant were considered. Each record is a datapoint for comparison. A minimum gap of 10 seconds was maintained between consecutive cough samples. 10 seconds is enough to trigger the cough receptors. The multiple recordings from the same participant helped in assessing the reliability of the device.

**Figure 1.**
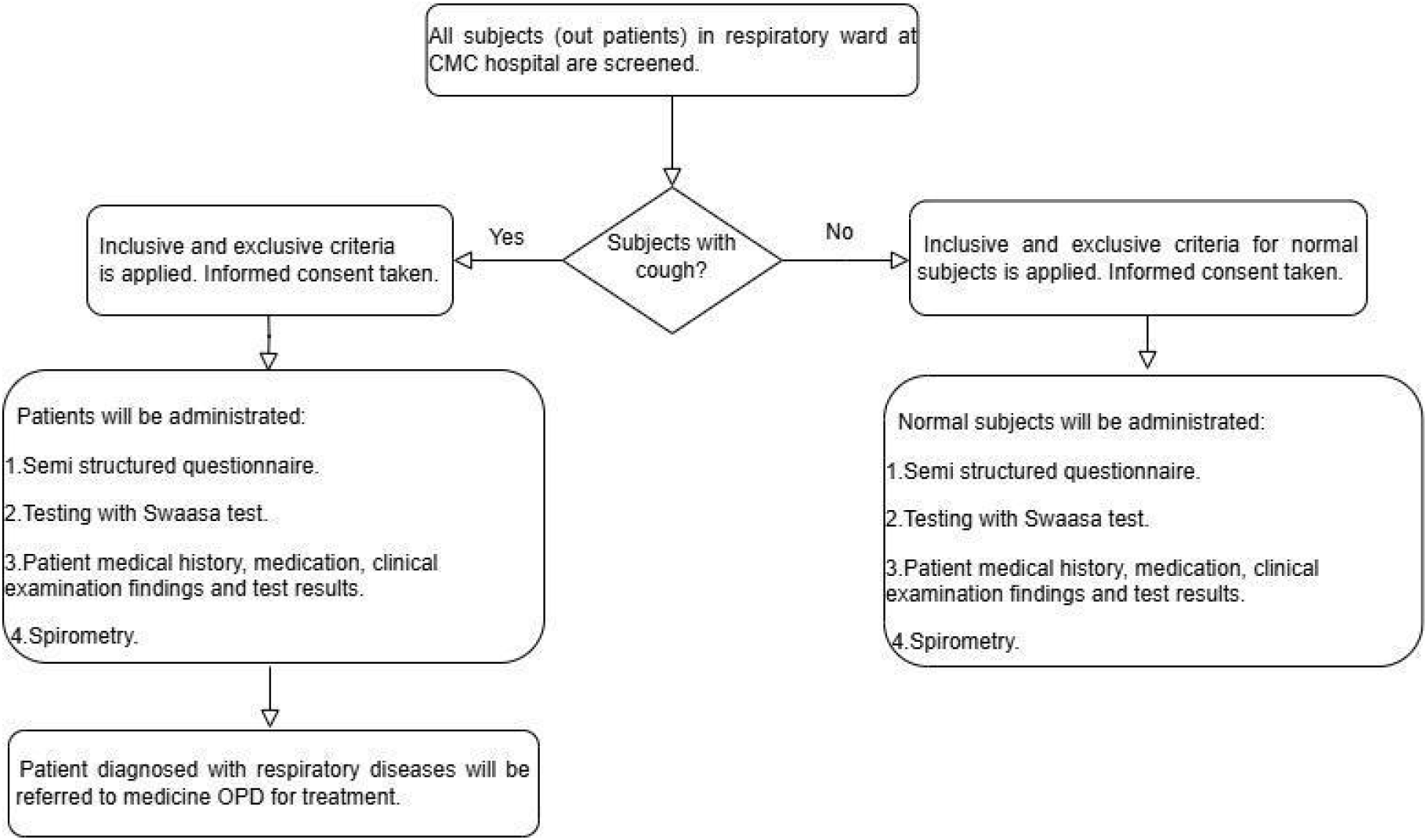
Data collection process

Spirometry is done according to American Thoracic Society guidelines 2019 (24). Abnormal spirometry was classified as obstruction or restriction based on FEV/FVC ratio. Those with isolated MMEF reductions were categorized separately. For calculation of accuracy measures, clinical diagnosis as concluded by the physician based on history, examination findings, spirometry and relevant radiological investigations was considered as the gold standard (ground truth). The procedure was double blinded to avoid bias. i.e. neither the subjects, therapists nor the doctor(s), were provided with the Swaasa report.

### Sample size

For the sample size calculation, based on the assumption that the device could present a sensitivity and specificity of 80% with an error of 5% for 95% CI in the detection of respiratory conditions, and a prevalence of the same of 25%, a sample size of 990 was found to be adequate for validation. However, to ensure rigor and improve the accuracy of the estimates, a larger sample size of 1,300 was estimated for a sensitivity/specificity of 90%. We would take one-fourth of this sample size to have an additional derivation cohort. Considering this and also to account for patients with incomplete data and those unable to do spirometry, we aimed to include a total of 1,800 participants in the study. Among the participants, 38% normal controls while the remaining were planned to be evenly distributed among four disease categories. (Asthma, COPD, ILD, Bronchiectasis). The normal subjects recruited in the study are the relatives’ accompanying patients to the outpatient clinic. These normal subjects are completely healthy subjects without symptoms. In the final analysis, a total of 1917 patients were available for validation.

### Study design

The study objective was to validate the Swaasa AI platform in a real-world setting of patients diagnosed with respiratory conditions. This explorative study had a monocentric, statistical methodology framework. It was conducted on the patients of the Respiratory and Pulmonary medicine departments of the Christian Medical College (CMC), Vellore.

The main objectives of the statistical validation exercise were to

(1) estimate the accuracy of Swaasa AI platform in detecting risk as normal vs abnormal, (2) estimate the accuracy of Swaasa AI platform in detecting respiratory disease pattern and severity (3) estimate the accuracy of the Swaasa AI platform in detecting disease conditions for Asthma, COPD, ILD and Bronchiectasis

The validation study included data from a prospective study, adopting a diagnostic accuracy study design. This process has been performed using 2 plans. Plan 1 was employed to assess risk (respiratory condition as normal / abnormal), pattern, and its severity, whereas plan 2 was applied for disease classification. These plans refer to the different ways in which the data was collected and organized for training and testing the AI model. In both the plans it was ensured that there was no overlap of subject data between the derivation and validation cohort.

The model used for validation including its input and output is illustrated in figure 2. The details about the pre-processing techniques, different architectures in finalizing the models are explained in our previous work (23, 26–27). The uniqueness of models lies in decoding the cough signatures for various conditions.

**Figure 2.**
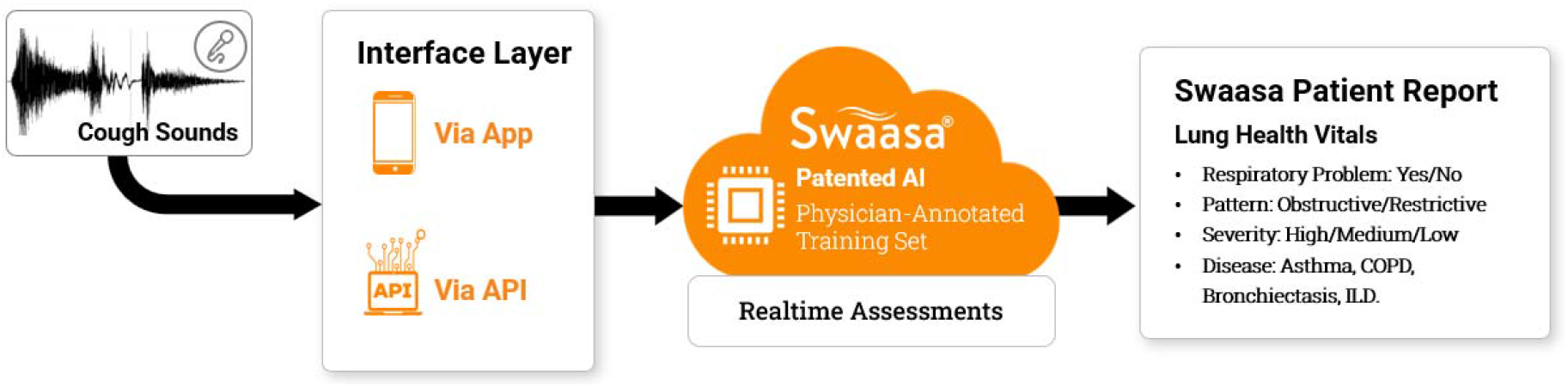
A flowchart highlighting the steps of Swaasa AI Platform

Labelling of respiratory conditions from the collected samples were performed by clinicians and were used as gold standard or ground truth annotation to assess the accuracy of the AI platform. The accuracy of binary and multi-class disease outcomes was assessed using accuracy rate, kappa statistics and diagnostic accuracy measures (Sensitivity & Specificity). The analysis is based on two distinct plans, Plan1 and Plan2, each designed to validate specific models.

Plan 1: The data from Plan 1 was utilized to validate models related to Risk, Spirometry Pattern, Pattern Severity. Among 1917 subjects, 450 were selected for derivation, and this dataset was incorporated into the existing training dataset. The remaining 1467 subjects are dedicated to the validation of the models mentioned above. (Plan 1 is used to predict Risk, Spirometry Pattern, Pattern Severity)

Plan 2: Plan 2 was focused on validating disease models. From the pool of 1917 subjects, 515 were chosen for derivation, and this dataset was integrated into the existing training dataset. The remaining 1402 subjects were employed for validating the performance of the disease models. (Plan 2 is used to predict Disease only).

Figure S1 in **supplement 1** shows the data distribution by age and sex.

## RESULTS

### Participants Information

From every participant three cough audio recordings were collected. In plan1 from 1467 subjects 3006 cough recordings were collected and in plan2 from 1402 subjects 2832 cough recordings were collected. Table 1 in details the distribution of clinical and demographic variables of the subjects. In this study, demographic and clinical variables and other variables of interest were summarized using frequency tables.

**Table 1:** Distribution of clinical and demographic variables.

|  | Data available for validation |  |
| --- | --- | --- |
| Variables | n (%)<br>N=3006<br>(Plan 1) | n (%)<br>N=2832<br>(Plan 2) |
| Age group |  |  |
| 18 – 47 years | 1636 (54.4) | 1559 (55.0) |
| 48 – 63 years | 934 (31.1) | 877 (31.0) |
| 64 – 89 years | 436 (14.5) | 396 (14.0) |
| Sex |  |  |
| Male | 1862 (61.9) | 1730 (61.1) |
| Female | 1144 (38.1) | 1102 (38.9) |

### Comparison of swaasa results with the ground truths

To evaluate the diagnostic accuracy of Swaasa as a screening tool, confusion matrix statistics were employed. These statistics included accuracy, Kappa statistics, sensitivity, and specificity. Accuracy, denoting the proportion of correct predictions out of all predictions made, serves as a fundamental measure of overall model performance. Table S1 and Table S2 in **Supplement 1** illustrate the comparison of swaasa results with the ground truths. The Kappa statistics assess the agreement level between the predictive model and observed data. Sensitivity, or the true positive rate, quantifies Swaasa’s ability to correctly identify true positive cases. Specificity, or the true negative rate, measures the model’s capacity to accurately identify true negative cases (controls). These metrics collectively provide a comprehensive understanding of the model’s proficiency in classifying outcomes.

Swaasa AI platform predicts Risk, Spirometry Pattern, Pattern Severity and Disease type from cough audio recording.

#### Risk

It refers to the detection of the presence of an underlying respiratory problem. If any abnormalities are detected in a subject, Swaasa classifies the Risk as “Abnormal”; otherwise, it categorizes them as “Normal.”

#### Spirometry Pattern

The term pattern refers to the detected respiratory disease pattern. Swaasa classifies patterns as “Obstructive,” “Restrictive,” or “Normal” based on the features extracted from the input data. Those with MMEF reduction alone were categorized separately.

#### Pattern Severity

Pattern severity refers to the assessment of the severity of respiratory disease patterns mentioned above. The severity categories include “Normal”, “Mild”, “Moderate,” and “Severe.” The spirometry report values are used as the reference to generate ground truth data and to build the severity model. When the spirometry report says normal, the ground truth of the patient is considered to be Normal. The severity categories are mapped to the FEV1% predicted values such as: Mild (>70), Moderate (70-50), Severe (<50) (25).

#### Disease Model

The Disease model is focused on detecting various respiratory conditions. Swaasa outputs diseases as “Asthma,” “COPD,” “ILD,” “Bronchiectasis,” and “Normal.” In the case of Abnormal vs. Normal detection, all diseases, including multi-labelled values from the ground truth, are considered abnormal.

### Performance Metrics of the Swaasa AI Models

Results highlight Swaasa’s proficiency in distinguishing between normal and abnormal conditions under all major domains or conditions, indicating its promise as a screening tool. The crosstabulations Table S3, Table S4 and Table S5 in **Supplement 1** shows Swaasa’s reasonable potential in identification of specific respiratory diseases and their severity.

Confusion matrix statistics for risk, pattern, severity and disease with binary classification for complete data is in Table S6, at gender level is in Table S7, and at sub age level is in Table S8.

Confusion matrix statistics for risk, pattern, severity and disease with multi-class variables for complete data is in Table 2.

**Table 2:**
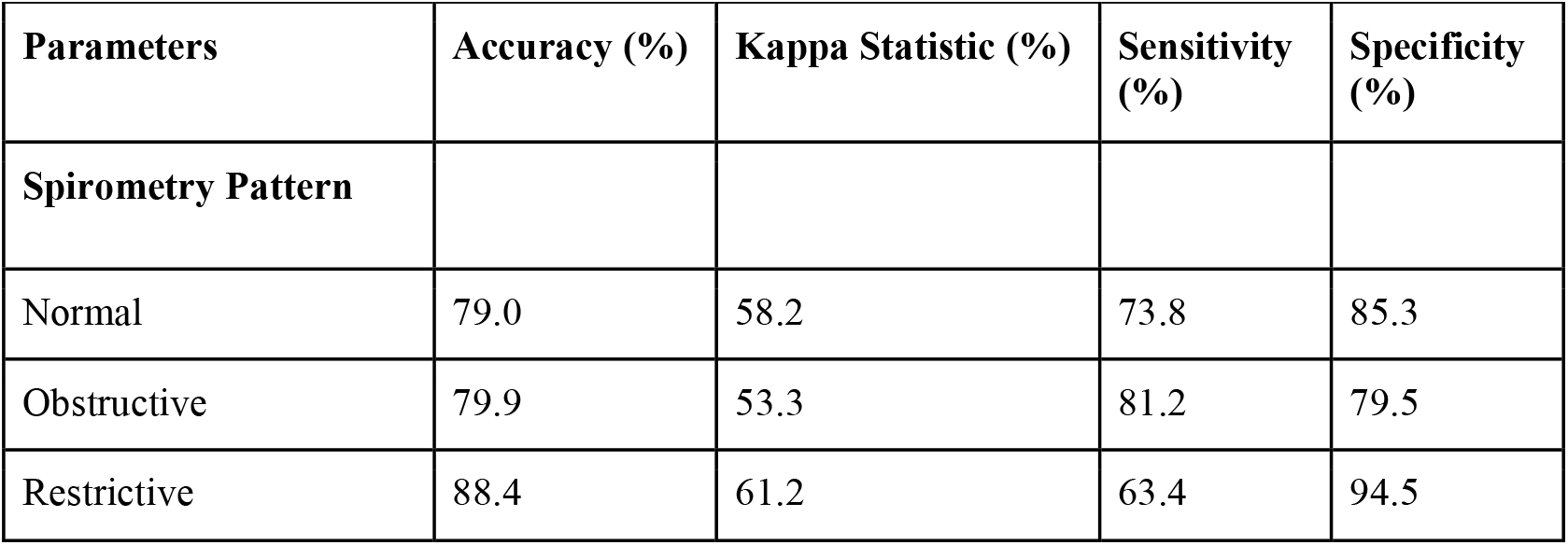
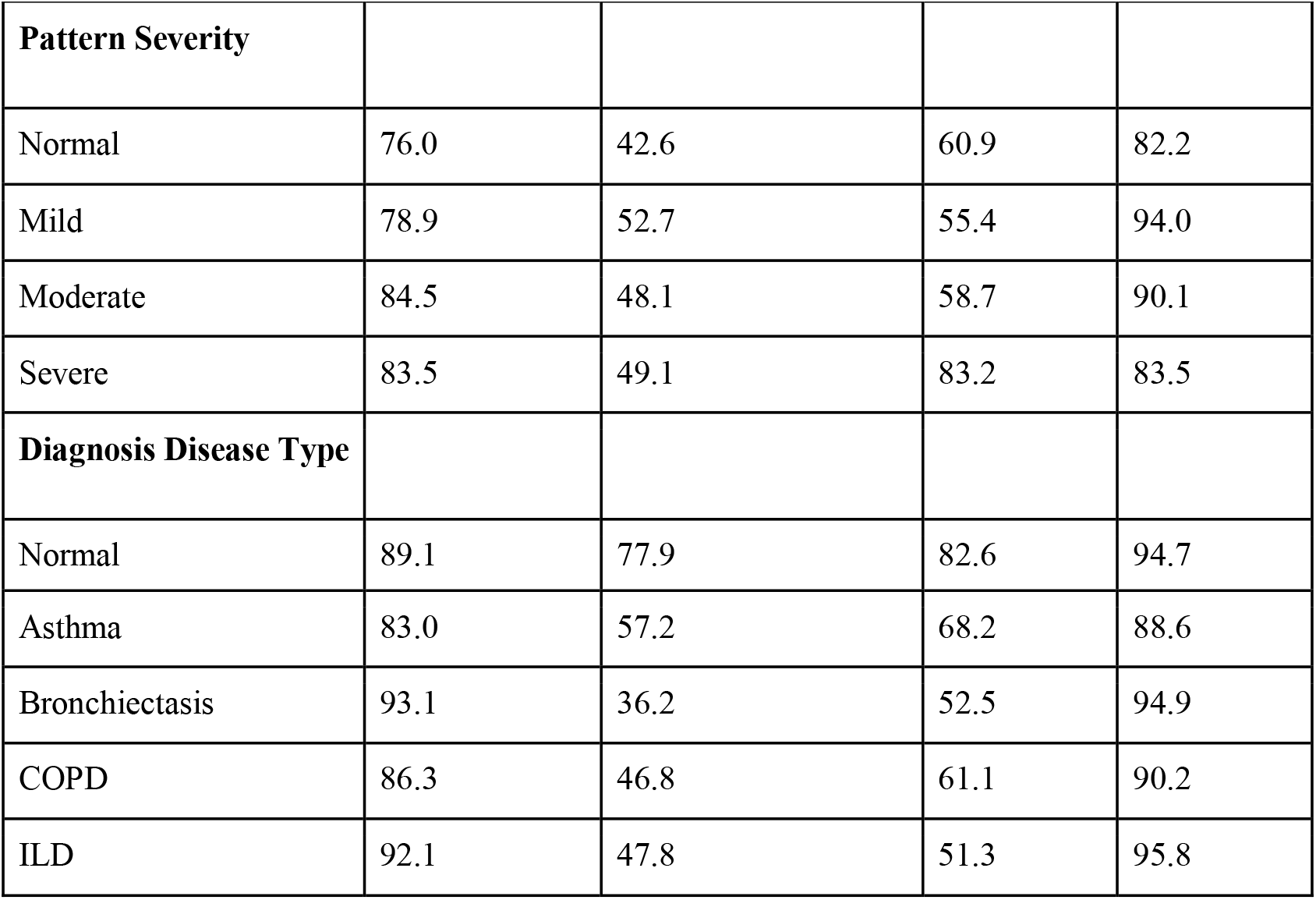
Confusion matrix statistics for spirometry pattern, spirometry severity and disease with multi-class variables.

| Parameters | Accuracy (%) | Kappa Statistic (%) | Sensitivity (%) | Specificity (%) |
| --- | --- | --- | --- | --- |
| **Spirometry Pattern** |  |  |  |  |
| Normal | 79.0 | 58.2 | 73.8 | 85.3 |
| Obstructive | 79.9 | 53.3 | 81.2 | 79.5 |
| Restrictive | 88.4 | 61.2 | 63.4 | 94.5 |

| **Pattern Severity** |  |  |  |  |
| --- | --- | --- | --- | --- |
| Normal | 76.0 | 42.6 | 60.9 | 82.2 |
| Mild | 78.9 | 52.7 | 55.4 | 94.0 |
| Moderate | 84.5 | 48.1 | 58.7 | 90.1 |
| Severe | 83.5 | 49.1 | 83.2 | 83.5 |
| **Diagnosis Disease Type** |  |  |  |  |
| Normal | 89.1 | 77.9 | 82.6 | 94.7 |
| Asthma | 83.0 | 57.2 | 68.2 | 88.6 |
| Bronchiectasis | 93.1 | 36.2 | 52.5 | 94.9 |
| COPD | 86.3 | 46.8 | 61.1 | 90.2 |
| ILD | 92.1 | 47.8 | 51.3 | 95.8 |

## Discussion

The Swaasa AI platform using artificial intelligence technology for screening respiratory diseases is not meant to replace or compete with the medical clinical diagnosis by any means. Instead, the proposed solution offers the following complementing use cases.

1. Addressing the shortage of testing facilities in isolated places. This is particularly useful in remote areas where Swaasa can act as a clinical decision assistance tool.
2. Maximizing time for medical experts by identifying the people at risk with respiratory issues.
3. Swaasa can be used as a low-cost screening tool, this is possible because our tests show that the device can identify respiratory conditions even in a non-spontaneous cough. The cost of using such an app-based solution would be significantly low, since it can be readily installed on smartphones using the existing internet connections, by many people simultaneously.
4. This device needs additional testing in multi-centric trials involving primary care healthcare and community settings.

Several studies have been conducted all over the world in the past to deploy information contained in the cough sound to detect and predict different disease outcomes such as Asthma, Pneumonia, COPD, bronchitis (10, 29-31). In recent times, due to the COVID-19 pandemic, there has been a tremendous boost in the use of ML/DL frameworks to determine the presence of SARS-CoV-2 infection using cough sample analysis. This is because coughing is one of the most prominent symptoms for diseases that primarily affect the respiratory system. Numerous studies have shown that cough analysis can accurately predict COVID-19 (3,5).

The strong concordance between the Swaasa AI platform and spirometry highlights the potential of AI-driven diagnostics in primary healthcare. Its ability to accurately identify obstructive lung diseases, such as COPD and asthma, suggests that cough sound analysis could be a practical alternative to traditional diagnostic methods, especially in regions where access to spirometry is limited.

The Swaasa AI platform offers several advantages over conventional diagnostic methods. Its non-invasive nature eliminates the need for specialized equipment and invasive procedures, making it suitable for a wide range of healthcare settings. Moreover, the platform’s cost-effectiveness and minimal training requirements make it an attractive option for rural healthcare centers, where trained personnel and diagnostic tools are often scarce.

## Limitations

Despite the rigorous methodology applied in this study, several limitations should be acknowledged in this study. The classification of respiratory diseases relied on clinician assessment, incorporating multidisciplinary discussions, guidelines-based approaches (such as GOLD and GINA), and radiological investigations. However, overlapping features of conditions like asthma and COPD or cough contributions from other factors (such as GERD) could not be clearly distinguished, introducing potential confounding variables. Similarly, a patient with COPD could have coexisting bronchiectasis. While spirometry abnormality was mapped to FEV1 categories for severity analysis, this may not be the most appropriate measure for diseases like asthma or ILD. In ILD, more relevant markers such as FVC or DLCO would offer a clearer representation of disease severity. The study’s device did not evaluate all potential respiratory diseases like cough due to post nasal drip, GERD, respiratory infections, tumors, etc. Further studies specifically designed for these are required. Further this is a study based in a tertiary care hospital. Further muti-centre studies based in primary health care or community settings may be required before this device is considered for clinical use.

## Conclusion

This paper presents a deployable AI-based preliminary diagnosis tool for screening respiratory diseases like Asthma, COPD, ILD and Bronchiectasis using cough sound as a maker. The core idea of the tool is inspired by our independent prior studies that show cough can be used as a medium for diagnosis of a variety of respiratory diseases using AI. We note that the way pathological condition affects the respiratory system is substantially unique and varies from respiratory condition to condition, cough associated with it is likely to have unique latent features as well. We validated the idea further by the visualization of latent features in cough of patients with different respiratory diseases like Asthma, COPD, ILD, bronchiectasis as well as healthy people coughs. Building on the insights from the medical domain knowledge, we developed an AI-platform for the cough-based diagnosis of respiratory conditions, named Swaasa AI platform. The results show that the device can diagnose respiratory conditions without needing clinical infrastructure.

## Data Availability

All data produced in the present study are available upon reasonable request to the authors

## Data Availability

Due to the nature of this research, participants of this study did not agree for their data to be shared publicly. However, the detailed analysis can be shared by the author “NRS” upon reasonable request.

## Ethical clearance

The study was registered under Clinical Trials Registry-India (CTRI/2021/04/032742) and was begun after getting the approval [Ethics Approval number - IRB Min. No. 13566 (DIAGNO)] from the CMC-IRB (Institutional Review Board). And all research was performed in accordance with relevant guidelines/regulations.

## Author Contributions

STC and BT defined study protocols, including study design and methodology. NRS conceptualized the idea of using cough sounds for screening and diagnosing respiratory diseases. GR performed literature review and data analysis. BM, HVR, SDP and NKRB were involved in device development. VY and MJ created value propositions for the device. SS assisted in executing the project at Christian Medical College by providing all the resources and extending research capabilities. CG and GR performed data analysis, sample size estimation and result analysis. KLPK, SS, NJ, VSP, ST and SV provided subject matter expertise. GR and JG wrote the manuscript. All the authors provided intellectual inputs and helped in preparing the manuscript.

## Acknowledgements

This study is supported by Biotechnology Industry Research Assistance Council (BIRAC) [grant number - BT/BIPP1335/BIPP-49/20]. We would also like to acknowledge the team from Christian Medical College, Vellore, India for all the support provided.

## Patient and Public Involvement

Patients or the public WERE NOT involved in the design, or conduct, or reporting, or dissemination plans of our research.

