## Supplement_1 for "CLINICAL VALIDATION OF SWAASA ARTIFICIAL INTELLIGENCE PLATFORM USING COUGH SOUNDS FOR SCREENING AND DIAGNOSIS OF RESPIRATORY DISEASES"

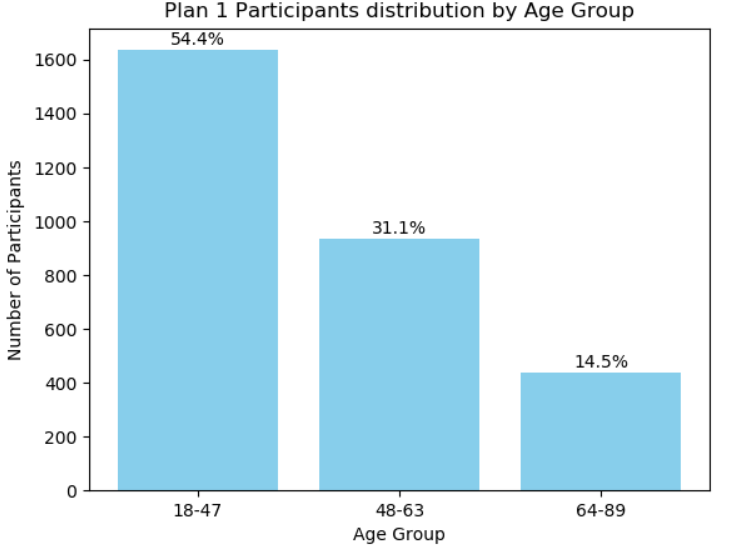

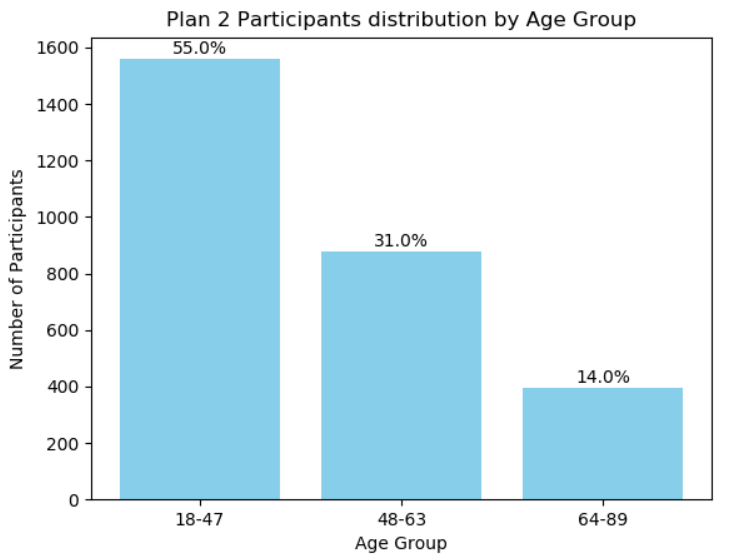

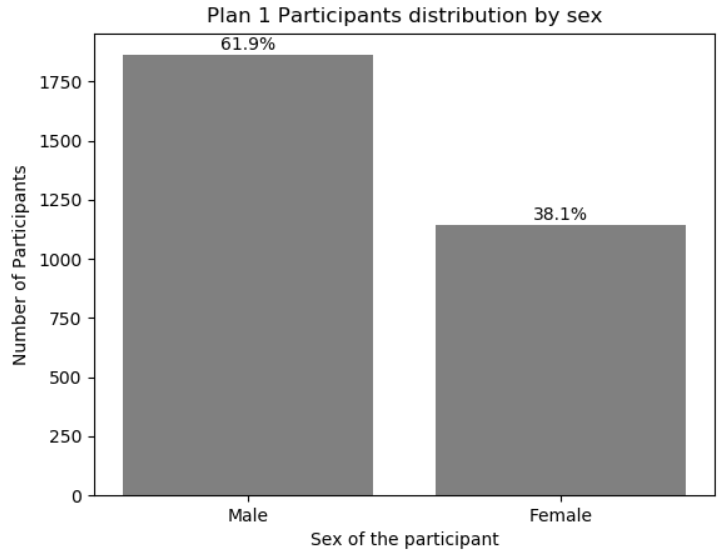

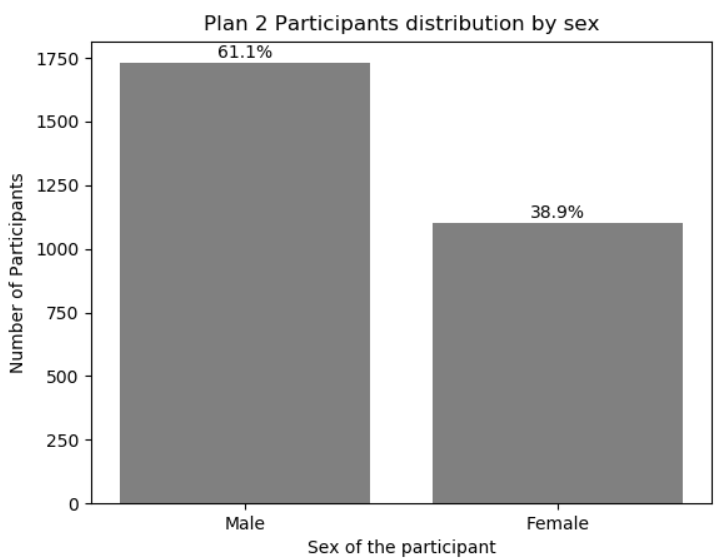

Figure S1: Data distribution in Plan 1 and Plan 2

Table S1: Frequencies for the Ground Truth and Swaasa risk, Spirometry pattern variables with binary classification

| Outcomes | Ground truth (n%) | Swaasa (n%) |
| --- | --- | --- |
| Risk |  |  |
| Normal | 827(27.5) | 835(27.8) |
| Abnormal | 2179(72.5) | 2171(72.2) |
| Total | 3006(100.0) | 3006(100.0) |
| Spirometry pattern |  |  |
| Normal | 1486(49.4) | 1377(45.8) |
| Abnormal | 1214(40.4) | 1639(54.2) |
| Missing | 306(10.2) | NA |
| Total | 3006(100.0) | 3006(100.0) |

Table S2: Frequencies for Ground Truth and Swaasa and disease variables with multiple classes

| Outcomes | Ground truth (n%) | Swaasa (n%) |
| --- | --- | --- |
| Diagnosis disease type |  |  |
| Normal | 852(30.1) | 857(30.3) |
| Asthma | 510(18.0) | 746(26.3) |
| Bronchiectasis | 80(2.8) | 286(10.1) |
| COPD | 252(9.0) | 577(20.4) |
| ILD | 156(5.5) | 365(12.9) |
| Multi - labelled | 845(29.8) | NA |
| Missing | 137(4.8) | 1(0.0) |
| Total | 2832(100.0) | 2832(100.0) |

To evaluate the diagnostic accuracy of Swaasa^®^ as a screening tool, confusion matrix statistics were employed. These statistics included accuracy, Kappa statistic, sensitivity, and specificity. Accuracy, denoting the proportion of correct predictions out of all predictions made, serves as a fundamental measure of overall model performance. The Kappa statistic assesses the agreement level between the predictive model and observed data. Sensitivity, or the true positive rate, quantifies Swaasa^’^ s ability to correctly identify true positive cases.  Specificity, or the true negative rate, measures the model's capacity to accurately identify true negative cases (controls). These metrics collectively provide a comprehensive understanding of the model's proficiency in classifying outcomes. All the metrics were derived using the cross tabulations etable 1, etable 2 and etable 3.

Table S3. Crosstabulation of Swaasa^®^ risk and underlying respiratory problem with binary classification along with column percentages

| Swaasa^®^ Risk | Underlying Respiratory problem | | |
| --- | --- | --- | --- |
|  | Normal | Abnormal | Total |
| Normal | 667 (80.7) | 168 (7.7) | 835 (27.8) |
| Abnormal | 160 (19.3) | 2011 (92.3) | 2171 (72.2) |
| Total | 827 (100) | 2179 (100) | 3006 (100) |

Table S4. Crosstabulation of Swaasa^®^ and ground truth pattern severity with multiple classes along with column percentages

| Swaasa^®^ Pattern Severity |  | GT Pattern Severity | | | | | |
| --- | --- | --- | --- | --- | --- | --- | --- |
|  |  | Normal | Mild | Moderate | Severe | Missing | Total |
| Normal |  | 479 (60.9) | 256 (24.2) | 67 (13.9) | 18 (4.8) | 26 (8.5) | 846 (28.1) |
| Mild |  | 68 (8.7) | 586 (55.4) | 16 (3.3) | 14 (3.7) | 20 (6.5) | 704 (23.4) |
| Moderate |  | 109 (13.9) | 79 (7.5) | 283(58.7) | 31 (8.3) | 128 (41.8) | 630 (21.0) |
| Severe |  | 130 (16.5) | 137(12.9) | 116 (24.1) | 311 (83.2) | 132 (43.1) | 826 (27.5) |
| Total |  | 786 (100) | 1058 (100) | 482 (100) | 374 (100) | 306 (100) | 3006 (100) |

Table S5. Crosstabulation of Swaasa^®^ and ground truth disease type pattern with multiple classes along with column percentages

| Swaasa^®^  Disease | Diagnosis Disease Type | | | | | | | |
| --- | --- | --- | --- | --- | --- | --- | --- | --- |
|  | Normal | Asthma | Bronchiectasis | COPD | ILD | Multi  labelled | Missing | Total |
| Normal | 703 (82.5) | 25 (4.9) | 8 (10.0) | 12 (4.8) | 8 (5.1) | 69 (8.2) | 32 (23.4) | 857 (30.3) |
| Asthma | 98 (11.5) | 348 (68.2) | 10 (12.5) | 28 (11.1) | 16 (10.3) | 226 (26.7) | 20 (14.6) | 746 (26.3) |
| Bronchiectasis | 13 (1.5) | 18 (3.5) | 42 (52.5) | 35 (13.9) | 24 (15.4) | 139 (16.4) | 15 (10.9) | 286 (10.1) |
| COPD | 16 (1.9) | 100 (19.6) | 12 (15.0) | 154 (61.1) | 28 (17.9) | 226 (26.7) | 41 (29.9) | 577 (20.4) |
| ILD | 21 (2.5) | 19 (3.7) | 8 (10.0) | 23 (9.1) | 80 (51.3) | 185 (21.9) | 29 (21.2) | 365 (12.9) |
| Missing | 1 (0.1) | 0 (0.0) | 0 (0.0) | 0 (0.0) | 0 (0.0) | 0 (0.0) | 0 (0.0) | 1 (0.0) |
| Total | 852 (100) | 510 (100) | 80 (100) | 252 (100) | 156 (100) | 845 (100) | 137 (100) | 2832 (100) |

Table S6: Confusion matrix statistics for disease identification and spirometry abnormality with binary classification (differentiating normal and abnormal).

| **Parameters** | **Accuracy (%)** | **Kappa Statistic (%)** | **Sensitivity (%)** | **Specificity (%)** |
| --- | --- | --- | --- | --- |
| Risk | 89.1 | 72.7 | 92.3 | 80.7 |
| Spirometry Pattern | 79.0 | 58.2 | 85.3 | 73.8 |
| Pattern Severity | 76.0 | 42.6 | 82.2 | 60.9 |
| Disease Type | 90.0 | 76.6 | 93.4 | 82.6 |

Table S7: Confusion matrix statistics for risk and pattern with binary classification at gender level.

| **Parameters** | **Accuracy (%)** | | **Kappa Statistic (%)** | |
| --- | --- | --- | --- | --- |
|  | **Male** | **Female** | **Male** | **Female** |
| Risk | 88.2 | 90.6 | 68.6 | 78.2 |
| Spirometry pattern | 80.5 | 76.5 | 61.2 | 52.6 |

Table S8: Confusion matrix statistics for risk and pattern with binary classification at age level.

| **Parameters** | **Accuracy (%)** | | | **Kappa Statistic (%)** | | |
| --- | --- | --- | --- | --- | --- | --- |
|  | **18-47 yrs** | **48-63 yrs** | **64-89 yrs** | **18-47 yrs** | **48-63 yrs** | **64-89 yrs** |
| Risk | 85.6 | 92.2 | 95.6 | 69.5 | 73.2 | 73.0 |
| Spirometry  Pattern | 77.2 | 79.8 | 84.5 | 52.3 | 59.2 | 61.2 |
